# Development and validation of nomograms for predicting the prognosis of early and late recurrence of advanced gastric cancer after radical surgery

**DOI:** 10.1101/2023.09.01.23294959

**Authors:** Chenming Liu, Jialiang Lu, Liang An

**Affiliations:** Department of general surgery, Shaoxing People’s Hospital, Shaoxing, Zhejiang province, China; Zhejiang University School of Medicine, Hangzhou, Zhejiang province, China; School of Medicine, Shaoxing University, Shaoxing, Zhejiang province, China; Department of gastrointestinal surgery, Shaoxing People’s Hospital, Shaoxing, Zhejiang province, China

**Keywords:** Gastric cancer, Gastrectomy, Recurrence, Nomogram, Prediction

## Abstract

**Objective:** In this study, we aimed to explore the risk factors influencing post recurrence survival (PRS) of early recurrence (ER) and late recurrence (LR) in stage advanced gastric cancer (AGC) patients after radical surgery, respectively, and to develop predictive models in turn.

**Methods:** Medical records of 192 AGC patients who recurred after radical gastrectomy were retrospectively reviewed. They were randomly divided into the training and validation set at a ratio of 2:1. Nomograms were built based on risk factors influencing PRS of ER and LR explored by Cox regression analyses, respectively. Concordance index (C-index) values and calibration curves were used to evaluate predictive power of nomograms.

**Results:** Body mass index < 18.5 kg/m^2^, prealbumin level < 70.1 mg/l, positive lymph nodes ratio ≥ 0.486 and palliative treatment after recurrence were independent risk factors for the prognosis of ER. In contrast, prealbumin level < 170.1 mg/l, CEA ≥ 18.32 μg/l, tumor diameter ≥ 5.5 cm and palliative treatment after recurrence were independent risk factors for the prognosis of LR. The C-index value was 0.801 and 0.772 for ER and LR in the training set, respectively. The calibration curves of validation set showed a C-index value of 0.744 and 0.676 for ER and LR, respectively.

**Conclusions:** Nomograms which were constructed to predict the prognosis of ER and LR of AGC after surgery showed great predictive power and could provide reference for clinicians’ treatment strategies to some extent.

## Introduction

Gastric cancer (GC) is the fifth most common malignancy and the fourth leading cause of cancer-related death worldwide.^1,2^ Although a variety of treatment methods have been gradually introduced into clinical practice in recent years, the prognosis of GC is still unsatisfactory and its five-year survival is no more than 20%.^3^ Radical gastrectomy is still the mainstream treatment strategy for patients with GC.^4^ More than half of GC patients experience recurrence within two years after surgery.^5,6^ Therefore, it is of great use to actively explore the prediction model accurately.

Previous studies have well elucidated the predictive role of certain tumor-specific factors during the recurrence of GC, especially early recurrence (ER), such as degree of tumor differentiation, tumor size, and perineural invasion.^7,8,9^ Furthermore, predictive models constructed based on these risk factors showed good predictive performance in practical clinical setting.^10,11^ But these studies included only patients with early GC or combined patients with early and advanced GC. AGC tends to have completely different characteristics from early GC due to its greater likeliness of invasion and metastasis. Predictors of prognosis after late recurrence (LR) of GC are not fully understood.

The objective of this study is to explore the risk factors influencing the prognosis of ER and LR in AGC patients after radical surgery respectively, and to build predictive models in turn, in order to timely spot high-risk patients and implement early intervention.

## Methods

### Study population

Medical records of patients with pathologically diagnosed AGC who underwent radical gastrectomy at our institution from 2016 to 2020 were retrospectively collected. Inclusion criteria were as follows: (1) age over 18 years at diagnosis; (2) neoadjuvant therapy before surgery was not performed; (3) recurrence was diagnosed by imaging or pathological examination after surgery. Patients who met the following criteria were excluded: (1) co-existing with other malignancies at diagnosis; (2) previous history of upper abdominal surgery; (3) R1 or R2 resection; (4) incomplete follow-up data. This study was approved by the Ethics Committee of our Institute and met the criteria of the Declaration of Helsinki.^12^ Random numbers generated by the computer divided the enrolled patients into the training set and the validation set in a 2:1 ratio. According to the latest surgical guidelines for GC in Japan, gastrectomy and D2 lymph node dissection were performed.^13^ The TNM staging of the tumor was based on the 8th edition of American Joint Committee on Cancer staging system.^14^ Fluorouracil and platinum-based regimen (usually 3-weel cycles of capecitabine/S-1 and oxaliplatin) was recommended for chemotherapy after surgery, depending on the patient’s physical condition and willingness.^15,16^ The detailed flow chart was shown in

### Variables

Clinicopathological variables included demographic information, surgery-related variables, pathology-related variables, and relevant nutritional and inflammatory variables at and after recurrence. As previously reported, prognostic nutritional index (PNI) was assessed using the following formula: PNI = serum albumin level (g/L) + 0.005 × peripheral blood total lymphocyte count (per mm^3^).^17^ The positive lymph node ratio (PLNR) was calculated as the total number of metastatic lymph nodes/total number of lymph nodes. In order to maintain the objectivity of the data, the receiver operating characteristic (ROC) curve was used to determine the optimal cut-off value for most continuous variables, such as, age and albumin. For detailed information, the additional material were shown (**S1 Table and S2 Table**). For body mass index (BMI) and anemia, widely accepted definitions were used.

### Definition

The cut-off value for the discrimination of ER and LR was defined as one year, based on criteria generally accepted in previous studies.^11,18^ Post-recurrence survival (PRS) was defined as the time interval between initial diagnosis of tumor recurrence and death or 3 years after recurrence. Recurrence included loco-regional, peritoneal and distant lymph nodes, hematogenous and multiple metastases. Loco-regional recurrence was defined as tumor recurrence in situ or local lymph node recurrence. Distant metastases included metastasis of the tumor to other parenchymal organs such as the liver, lungs, bone, and distant lymph nodes such as the pleura, neck, armpit, and subclavian lymph nodes. Multiple metastases were defined as two or more metastatic sites.

### Follow-up

Patients were followed up every 2-3 months for the first two years after discharge and every 6 months thereafter until recurrence. Follow-up included blood tests, chest X-ray, abdominal computed tomography or magnetic resonance imaging, and endoscopic tissue biopsy if necessary. Recurrence diagnosed by regular surveillance was defined as those patients diagnosed by abdominal medical imaging and/or tumor bio-markers at an interval of 2-3 months or less on asymptomatic patients. Recurrence diagnosed by irregular surveillance was defined as those patients having an interval of more than 3 months when recurrences were diagnosed by medical imaging or tumor bio-markers, or because investigations were carried out for symptoms or other unrelated reasons.^19^ Post-recurrence treatment included palliative treatment, adjuvant chemotherapy and re-surgical intervention, which were selected according to the practical condition and willingness of patients.

### Statistical analysis

For continuous variables distributed normally in the whole cohort, the mean and standard deviation (SD) were calculated, and a t-test was used to assess differences between groups. Otherwise, the median and the interquartile range (IQR) were calculated, and compared using a Wilcoxon test. Categorical variables were described as frequency (%) and analyzed using a chi-square test or Fisher’s exact test. Kaplan-Meier method and log-rank test were used for survival analysis. Multivariate Cox regression analysis were used to determine the independent risk factors for PRS with ER and LR respectively. Nomograms were constructed based on these risk factors. The data sets were divided into two groups: the larger data set was used to develop the model, and the smaller data set was used to externally validate the built model. The model’s predictions were evaluated according to the concordance index (C-index) of 0-1 and 95% confidence interval (CI) and the area under the curve (AUC) of the ROC curve. The bootstrap method (frequency =1000) was used for internal validation, the C-index was calculated and the calibration curve was drawn. All statistical analysis and data processing were performed SPSS software (version 25.0, IBM Corp., Armonk, NY, USA) and R software (version 4.1.2; R Foundation for Statistical Computing). All tests were bilateral, and *P* < 0.05 was considered significant.

## Results

### Characteristics of clinicopathological variables

Among the patients with ER, 73.3% were male, 80.2% were pTNM stage III, and 78.2% received postoperative adjuvant chemotherapy. For the recurrence site, distant hematogenous metastasis was the most common (32.7%). For post-recurrence treatment, 42.6% received chemotherapy and 12.9% received re-surgery intervention. In contrast, for patients with LR, 76.9% were male, 70.3% were pTNM stage III, 87.9% received postoperative adjuvant chemotherapy, and for the recurrence site, distant metastasis and peritoneal metastasis were most common (33.0%). For post-recurrence treatment, 49.5% received chemotherapy and 20.9% received re-surgery intervention. The detailed clinicopathological characteristics of ER and LR were shown in **Table 1** and **2**, respectively. No obvious differences were found between training and validation set (all *P* > 0.05).

**Table 1.**
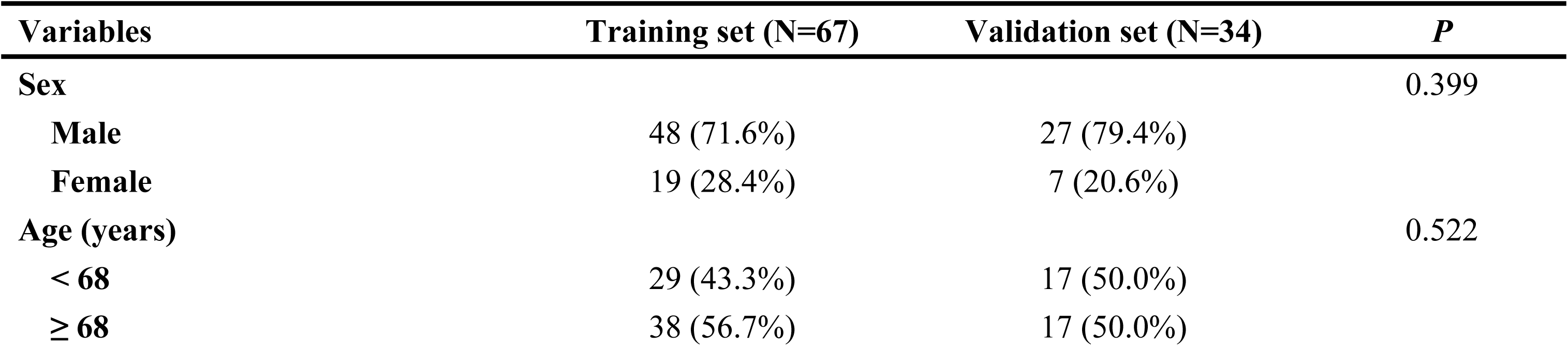
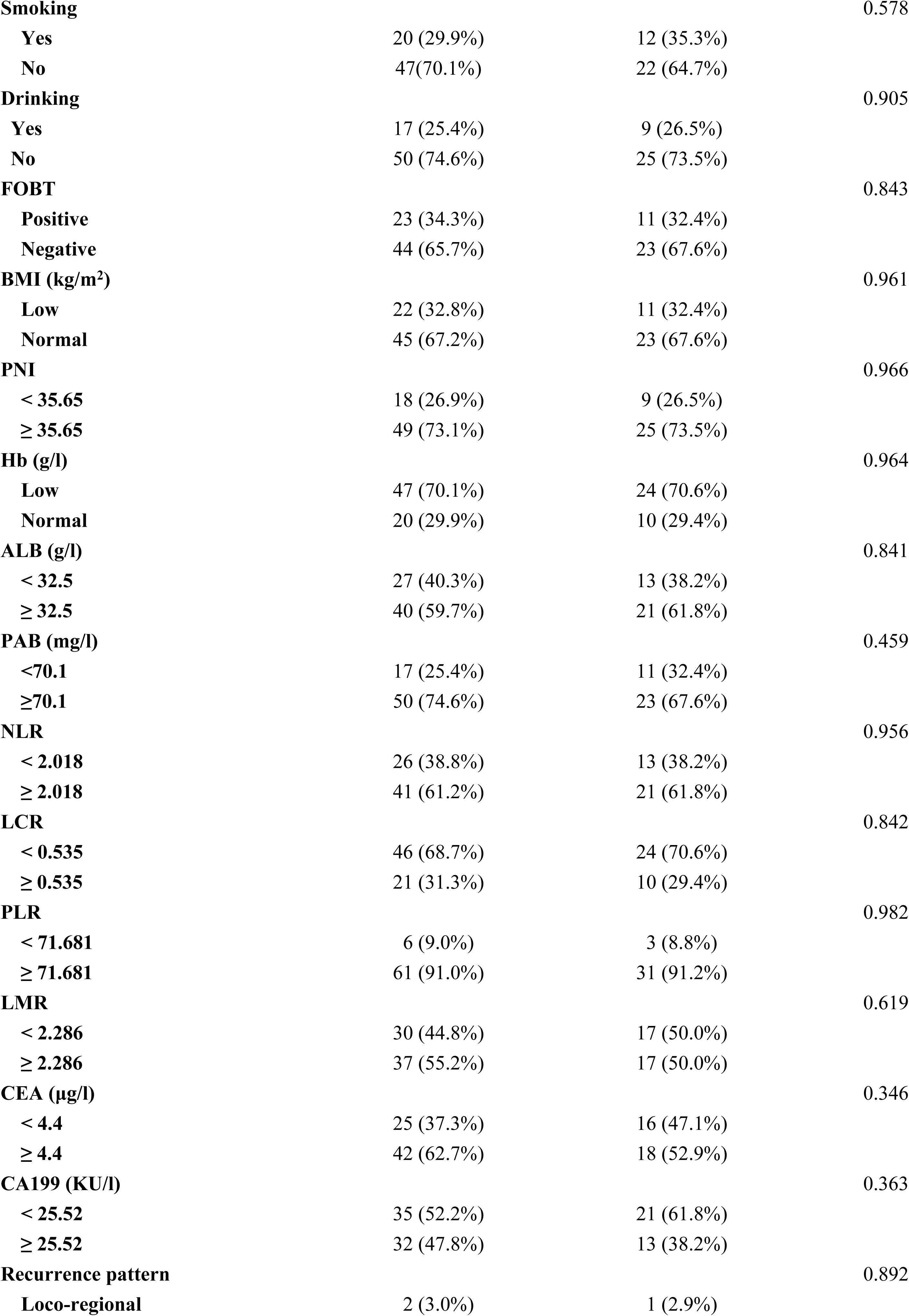
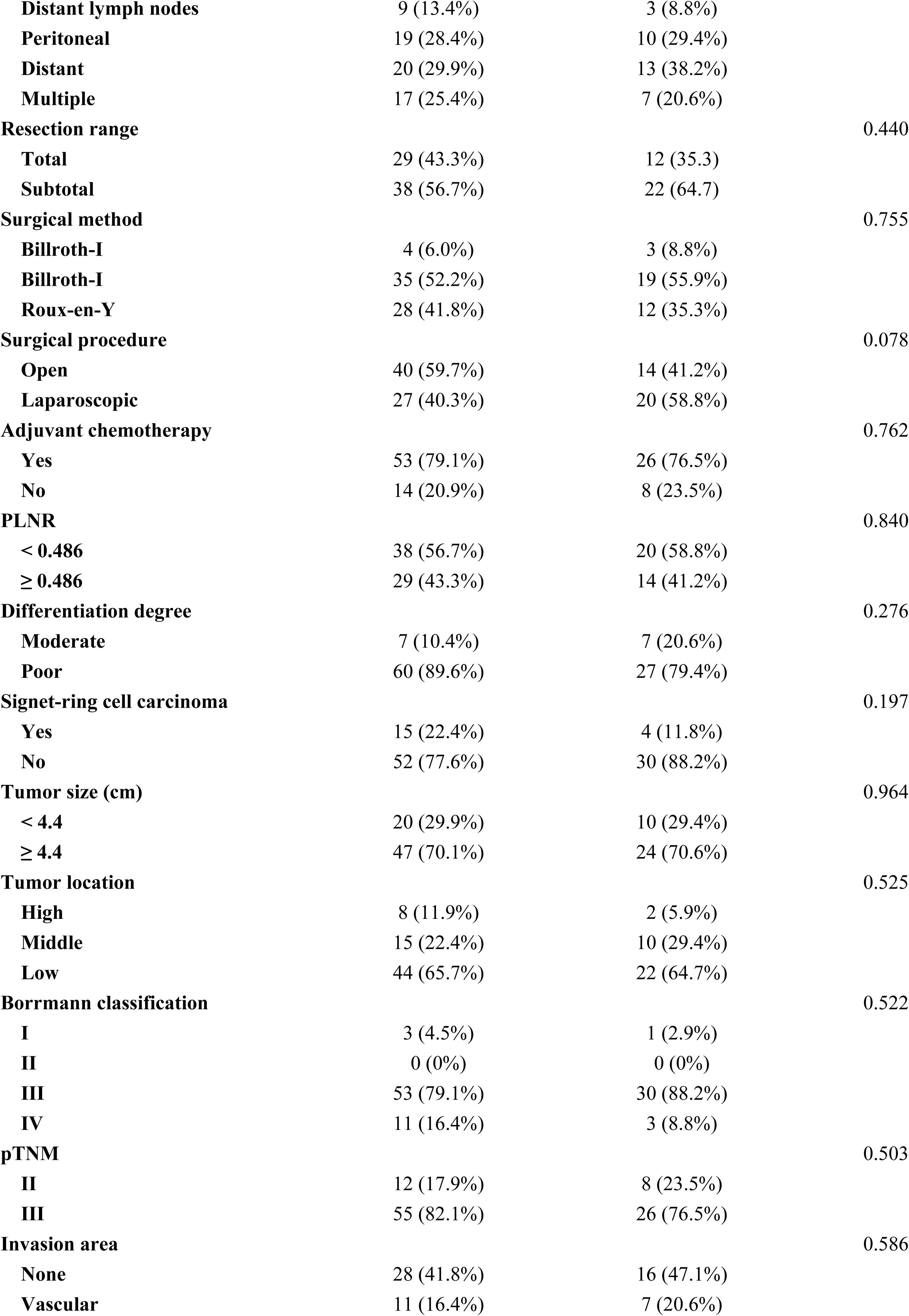
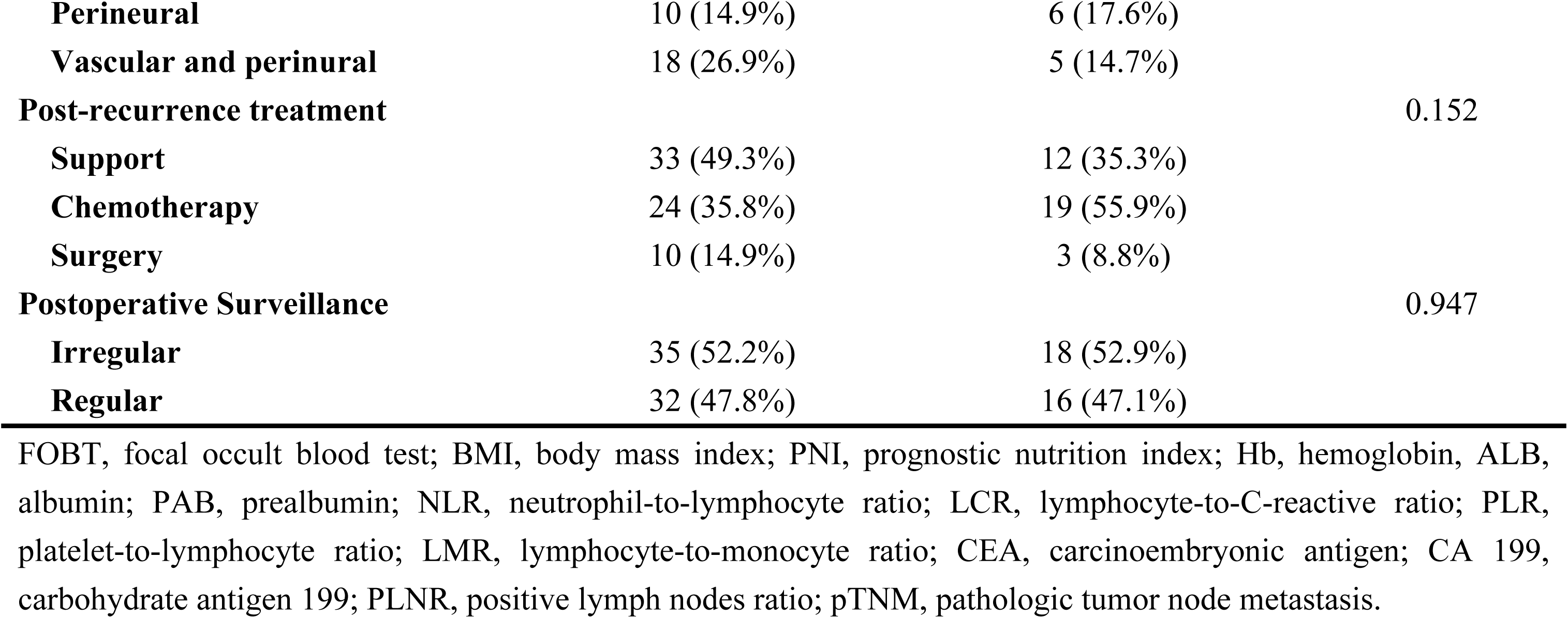
Comparison of characteristics between training and validation cohort in patients with early recurrence.

**Table 2.**
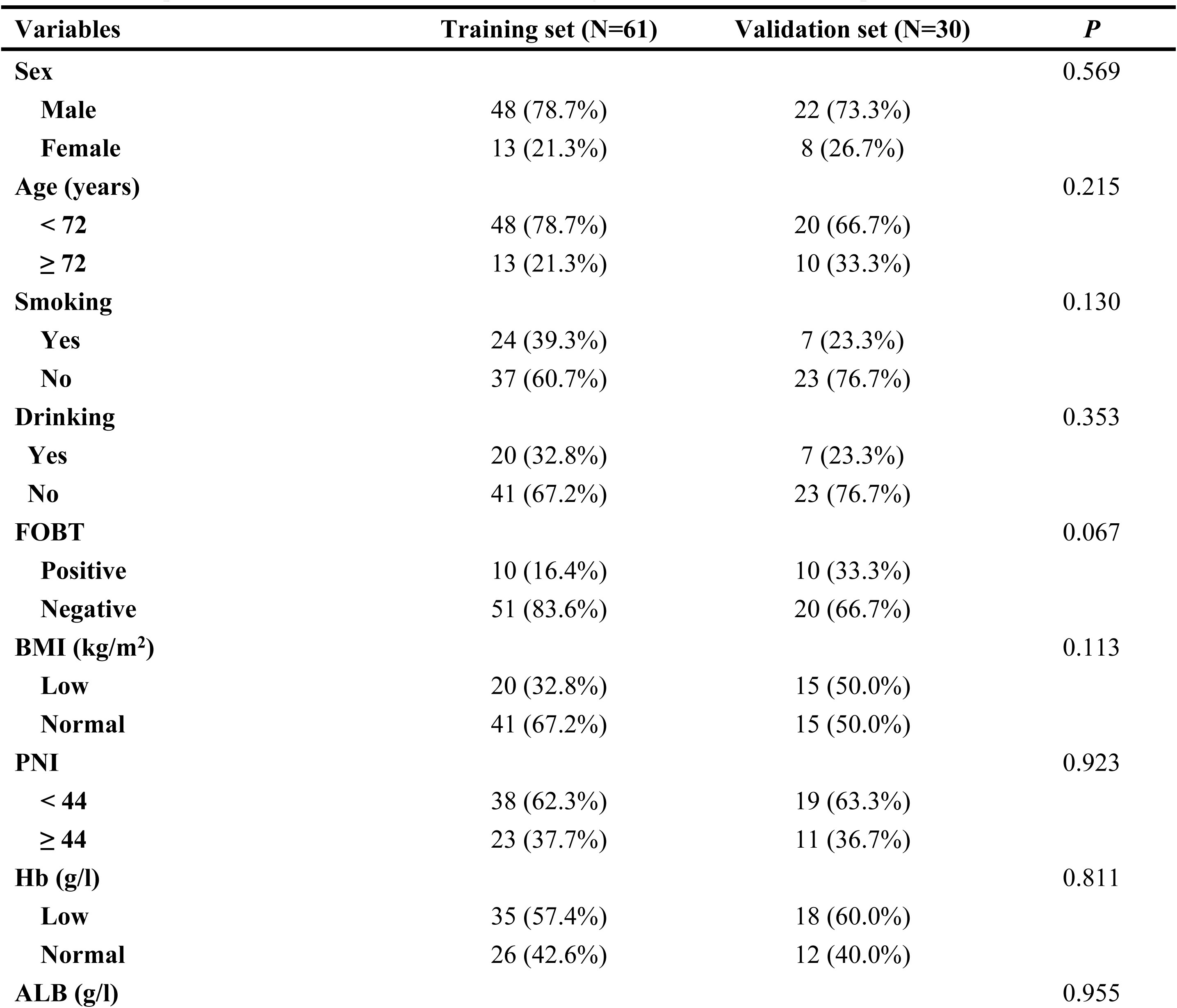
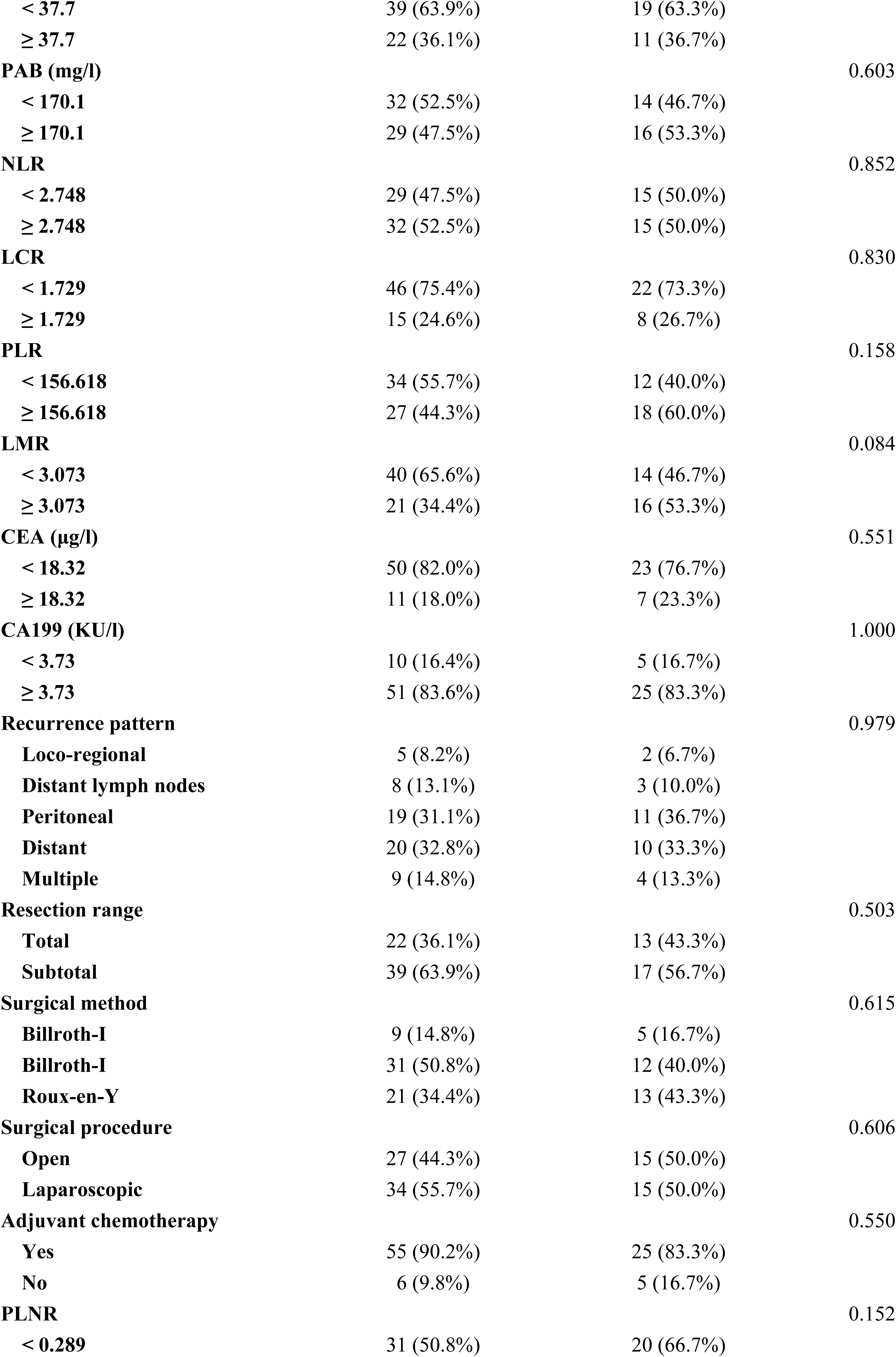
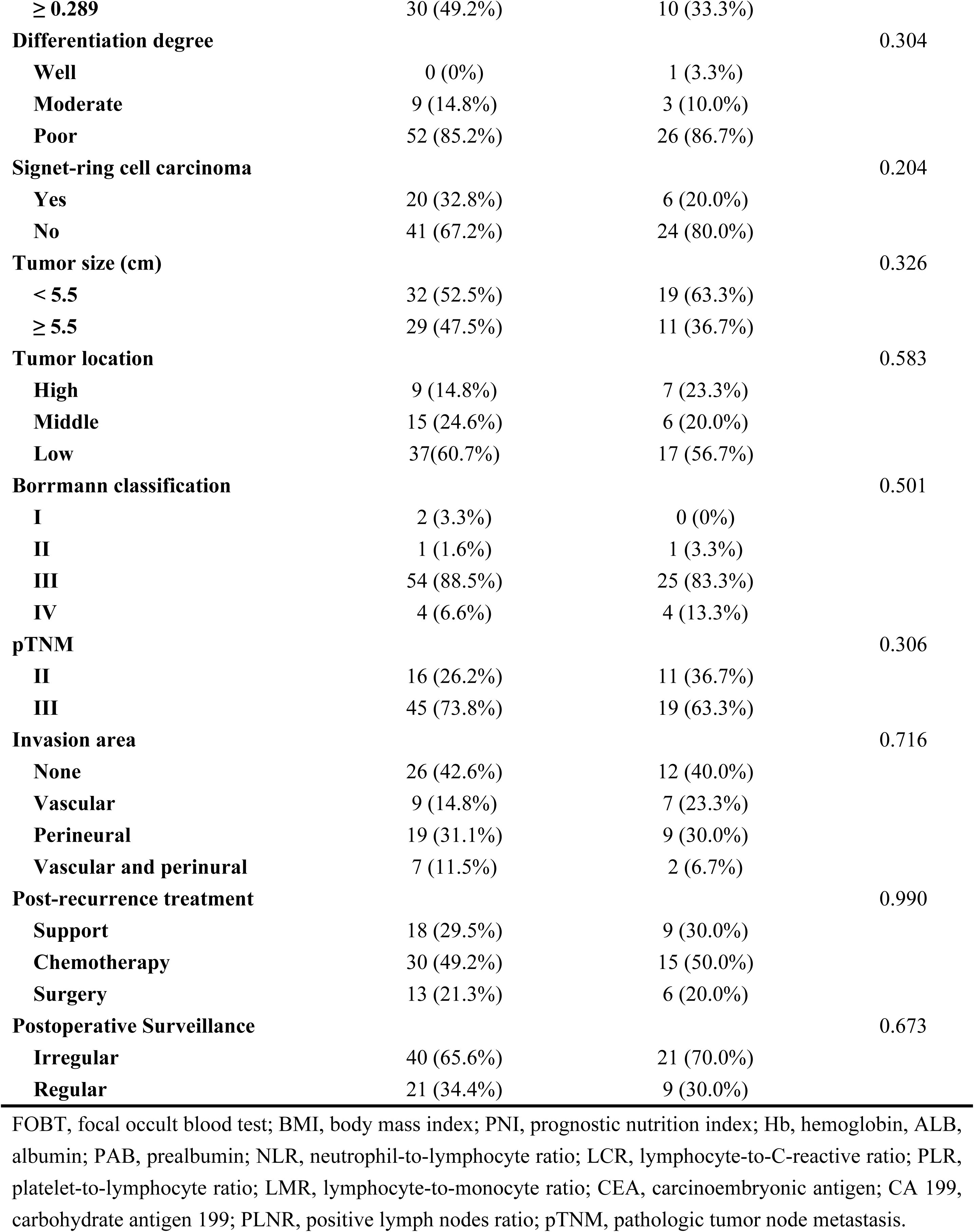
Comparison of characteristics between training and validation cohort in patients with late recurrence.

### Comparison of recurrence patterns

For patients with ER, the most common recurrence pattern was distant hematogenous metastasis (29.9%), followed by peritoneal metastasis (28.4%). To be slightly different, for patients with LR, the most common recurrence pattern was peritoneal metastasis (32.8%), followed by distant metastasis (31.3%). The recurrence patterns were similar between the two groups (all *P* > 0.05) (**Table 3**).

**Table 3.** Comparison of recurrence pattern between early and late recurrence.

| Recurrence pattern | Early recurrence (n=67) | Late recurrence (n=61) | *P* |
| --- | --- | --- | --- |
| Loco-regional | 2 (3.0%) | 5 (8.2%) | 0.365 |
| Distant lymph nodes | 9 (13.4%) | 8 (13.1%) | 0.958 |
| Peritoneal | 19 (28.4%) | 19 (31.1%) | 0.730 |
| Hematogenous | 20 (29.9%) | 20 (32.8%) | 0.720 |
| Multiple | 17 (25.4%) | 9 (14.8%) | 0.136 |

### Multivariate Cox regression analysis of patients with ER and LR

Multivariate Cox regression analysis showed that BMI < 18.5 kg/m^2^, prealbumin level < 70.1 mg/l, PLNR ≥ 0.486 and palliative treatment after recurrence were independent risk factors for the prognosis of ER (**Table 4**). In contrast, prealbumin level < 170.1 mg/l at recurrence, CEA ≥ 18.32 μg/l, tumor diameter ≥ 5.5 cm and palliative treatment after recurrence were independent risk factors for prognosis of LR (**Table 5**).

**Table 4.** Univariate and multivariate cox regression analysis of PRS in gastric cancer with early recurrence.

| Characteristics | Univariate analysis |  | Multivariate analysis |  |
| --- | --- | --- | --- | --- |
|  | HR (95% CI) | *P* | HR (95% CI) | *P* |
| Sex, female vs male | 1.334 (0.763-2.332) | 0.313 |  |  |
| Age (years), $\geq 68$ vs $< 68$ | 1.270 (0.755-2.137) | 0.368 | | |
| Smoking, yes vs no | 1.571 (0.913-2.703) | 0.103 |  |  |
| Drinking, yes vs no | 1.180 (0.663-2.098) | 0.574 |  |  |
| FOBT, positive vs negative | 1.118 (0.651-1.919) | 0.687 |  |  |
| BMI (kg/m<sup>2</sup>), normal vs low | 0.335 (0.193-0.581) | $< 0.001^*$ | 0.551 (0.303-0.999) | 0.049* |
| PNI, $\geq 35.65$ vs $< 35.65$ | 0.358 (0.201-0.638) | $< 0.001^*$ | | |
| Hb (g/l), normal vs low | 0.904 (0.515-1.589) | 0.727 |  |  |
| ALB (g/l), $\geq 32.5$ vs $< 32.5$ | 0.610 (0.363-1.026) | 0.062 | | |
| PAB (mg/l), $\geq 70.1$ vs $< 70.1$ | 0.326 (0.182-0.586) | $< 0.001^*$ | 0.289 (0.150-0.558) | $< 0.001^*$ |
| NLR, $\geq 2.018$ vs $< 2.018$ | 2.822 (1.619-4.916) | $< 0.001^*$ | | |
| LCR, $\geq 0.535$ vs $< 0.535$ | 0.525 (0.298-0.927) | 0.026* | | |
| PLR, $\geq 71.681$ vs $< 71.681$ | 3.304 (1.027-10.628) | 0.045* | | |
| LMR, $\geq 2.286$ vs $< 2.286$ | 0.507 (0.302-0.850) | 0.010* | | |
| CEA (μg/l), $\geq 4.4$ vs $< 4.4$ | 1.443 (0.833-2.502) | 0.191 | | |
| CA199 (KU/l), $\geq 25.52$ vs $< 25.52$ | 1.820 (1.081-3.062) | 0.024* | | |
| Recurrence pattern | Ref - loco-regional |  |  |  |
| Distant lymph nodes | 2.490 (0.305-20.307) | 0.394 |  |  |
| **Peritoneal** | 3.017 (0.401-22.712) | 0.284 |  |  |
| **Hematogenous** | 5.015 (0.667-37.684) | 0.117 |  |  |
| **Multiple** | 5.553 (0.728-42.391) | 0.098 |  |  |
| **Resection range, Subtotal vs total** | 0.979 (0.585-1.638) | 0.937 |  |  |
| **Surgical method** | Ref - Billroth-I |  |  |  |
| **Billroth-II** | 1.612 (0.489-5.318) | 0.433 |  |  |
| **Roux-en-Y** | 1.473 (0.442-4.909) | 0.528 |  |  |
| **Operative type, LAP vs OP** | 0.624 (0.366-1.063) | 0.082 |  |  |
| **Adjuvant chemotherapy, yes or no** | 1.027 (0.545-1.938) | 0.933 |  |  |
| **PLNR, $\geq 0.486$ vs $< 0.486$** | 2.842 (1.626-4.907) | $< 0.001^*$ | 2.879(1.607-5.159) | $< 0.001^*$ |
| **Differentiation, poor vs moderate** | 0.948 (0.407-2.211) | 0.902 |  |  |
| **Signet-ring cell carcinoma, yes vs no** | 0.947 (0.510-1.757) | 0.862 |  |  |
| **Tumor size (cm), $\geq 4.4$ vs $<4.4$** | 1.418 (0.795-2.530) | 0.237 |  |  |
| **Tumor location** | Ref- High |  |  |  |
| **Middle** | 1.265 (0.497-3.221) | 0.622 |  |  |
| **Low** | 1.729 (0.771-3.877) | 0.184 |  |  |
| **Borrmann classification** | Ref- I |  |  |  |
| **III** | 2.517 (0.609-10.395) | 0.202 |  |  |
| **IV** | 4.117 (0.891-19.017) | 0.070 |  |  |
| **pTNM, III vs II** | 1.674 (0.843-3.325) | 0.141 |  |  |
| **Invasion area** | Ref - none |  |  |  |
| **Peri-vascular** | 0.960 (0.458-2.011) | 0.914 |  |  |
| **Peri-neural** | 1.113 (0.500-2.481) | 0.793 |  |  |
| **Peri-vascular and neural** | 1.920 (1.016-3.631) | 0.045 |  |  |
| **Post-recurrence treatment** | Ref- palliative |  |  |  |
| **Adjuvant chemotherapy** | 0.456 (0.260-0.801) | 0.006* | 0.402 (0.214-0.755) | 0.005* |
| **Re-surgery intervention** | 0.332 (0.144-0.766) | 0.010* | 0.398 (0.171-0.925) | 0.032* |
| **Postoperative surveillance, no vs yes** | 2.184 (1.296-3.681) | 0.003* |  |  |

FOBT, focal occult blood test; BMI, body mass index; PNI, prognostic nutrition index; Hb, hemoglobin, ALB, albumin; PAB, prealbumin; NLR, neutrophil-to-lymphocyte ratio; LCR, lymphocyte-to-C-reactive ratio; PLR, platelet-to-lymphocyte ratio; LMR, lymphocyte-to-monocyte ratio; CEA, carcinoembryonic antigen; CA 199, carbohydrate antigen 199; LAP, laparoscopic; OP, open; PLNR, positive lymph nodes ratio; pTNM, pathologic tumor node metastasis.

**Table 5.** Univariate and multivariate cox regression analysis of PRS in gastric cancer with late recurrence.

| **Characteristics** | **Univariate analysis** |  | **Multivariate analysis** |  |
| --- | --- | --- | --- | --- |
|  | **HR (95 CI%)** | **P** | **HR (95 CI%)** | **P** |
| **Sex, female vs male** | 1.618 (0.855-3.062) | 0.140 |  |  |
| **Age (years), ≥ 72 vs < 72** | 1.638 (0.871-3.080) | 0.126 |  |  |
| **Smoking, yes vs no** | 0.849 (0.487-1.483) | 0.566 |  |  |
| **Drinking, yes vs no** | 1.014 (0.569-1.807) | 0.963 |  |  |
| **FOBT, positive vs negative** | 1.556 (0.776-3.120) | 0.213 |  |  |
| **BMI (kg/m<sup>2</sup>), normal vs low** | 0.404 (0.228-0.715) | 0.002* |  |  |
| **PNI, ≥ 44 vs < 44** | 0.454 (0.252-0.817) | 0.008* |  |  |
| **Hb (g/l), normal vs low** | 0.472 (0.269-0.827) | 0.009* |  |  |
| **ALB (g/l), ≥ 37.7 vs < 37.7** | 0.391 (0.215-0.711) | 0.002* |  |  |
| **PAB (mg/l), ≥ 170.1 vs < 170.1** | 0.354 (0.202-0.620) | < 0.001* | 0.426 (0.239-0.762) | 0.004* |
| **NLR, $\geq 2.748$ vs $< 2.748$** | 1.779 (1.021-3.101) | 0.042* |  |  |
| **LCR, $\geq 1.729$ vs $< 1.729$** | 0.492 (0.252-0.959) | 0.037* |  |  |
| **PLR, $\geq 156.618$ vs $< 156.618$** | 1.493 (0.864-2.578) | 0.151 |  |  |
| **LMR, $\geq 3.073$ vs $< 3.073$** | 0.669 (0.374-1.199) | 0.177 |  |  |
| **CEA ($\mu\text{g/l}$), $\geq 18.32$ vs $< 18.32$** | 2.469 (1.207-5.050) | 0.013* | 2.923 (1.369-6.240) | 0.006* |
| **CA199 (KU/l), $\geq 3.73$ vs $< 3.73$** | 1.869 (0.841-4.150) | 0.125 |  |  |
| **Recurrence pattern** | Ref - loco-regional |  |  |  |
| **Distant lymph nodes** | 0.416 (0.111-1.558) | 0.193 |  |  |
| **Peritoneal** | 1.263 (0.424-3.762) | 0.675 |  |  |
| **Hematogenous** | 1.080 (0.365-3.200) | 0.889 |  |  |
| **Multiple** | 1.813 (0.551-5.969) | 0.328 |  |  |
| **Resection range, Subtotal vs total** | 0.861 (0.493-1.505) | 0.600 |  |  |
| **Surgical method** | Ref- Billroth-I |  |  |  |
| **Billroth-II** | 1.327 (0.577-3.055) | 0.505 |  |  |
| **Roux-en-Y** | 1.468 (0.615-3.504) | 0.387 |  |  |
| **Operative type, LAP vs OP** | 1.101 (0.640-1.894) | 0.728 |  |  |
| **Adjuvant chemotherapy, yes or no** | 1.303 (0.512-3.312) | 0.578 |  |  |
| **PLNR, $\geq 0.289$ vs $< 0.289$** | 1.434 (0.833-2.467) | 0.194 |  |  |
| **Differentiation, poor vs moderate** | 1.189 (0.536-2.641) | 0.670 |  |  |
| **Signet-ring cell carcinoma, yes vs no** | 1.056 (0.597-1.870) | 0.851 |  |  |
| **Tumor size (cm), $\geq 5.5$ vs $< 5.5$** | 1.964 (1.131-3.413) | 0.017* | 2.497 (1.388-4.491) | 0.002* |
| **Tumor location** | Ref- high |  |  |  |
| **Middle** | 0.785 (0.324-1.902) | 0.592 |  |  |
| **Low** | 0.898 (0.412-1.958) | 0.787 |  |  |
| **Borrmann classification** | Ref- I |  |  |  |
| **II** | 0.413 (0.037-4.671) | 0.475 |  |  |
| **III** | 0.313 (0.073-1.348) | 0.119 |  |  |
| **IV** | 0.801 (0.144-4.465) | 0.800 |  |  |
| **pTNM, III vs II** | 1.152 (0.625-2.122) | 0.651 |  |  |
| **Invasion area** | Ref - none |  |  |  |
| **Peri-vascular** | 1.719 (0.782-3.778) | 0.177 |  |  |
| **Peri-neural** | 0.845 (0.442-1.613) | 0.609 |  |  |
| **Peri-vascular and neural** | 1.044 (0.421-2.587) | 0.926 |  |  |
| **Post-recurrence treatment** | Ref - palliative |  |  |  |
| **Adjuvant chemotherapy** | 0.376 (0.197-0.715) | 0.003* | 0.328 (0.159-0.674) | 0.002* |
| **Re-surgery intervention** | 0.395 (0.180-0.866) | 0.020* | 0.420 (0.184-0.955) | 0.039* |
| **Postoperative surveillance, no vs yes** | 2.160 (1.194-3.907) | 0.011* |  |  |

FOBT, focal occult blood test; BMI, body mass index; PNI, prognostic nutrition index; Hb, hemoglobin, ALB, albumin; PAB, prealbumin; NLR, neutrophil-to-lymphocyte ratio; LCR, lymphocyte-to-C-reactive ratio; PLR, platelet-to-lymphocyte ratio; LMR, lymphocyte-to-monocyte ratio; CEA, carcinoembryonic antigen; CA 199, carbohydrate antigen 199; LAP, laparoscopic; OP, open; PLNR, positive lymph nodes ratio; pTNM, pathologic tumor node metastasis.

### PRS for patients with ER and LR

Kaplan-Meier curves showed no significant difference in median PRS between patients with ER and LR (**Figure 2**). Subgroup survival analyses were then performed on the basis of prealbumin level and categories of post-recurrence treatment. The results showed that lower prealbumin level at recurrence had worse prognosis than higher prealbumin level (ER: 1 vs 6 months, *P* < 0.001; LR: 3 vs 13 months, *P* < 0.001) (**S1 Figure** and **S2 Figure)**. Compared with re-surgery intervention and adjuvant chemo-radiotherapy, patients who received palliative treatment after recurrence had worse PRS (for ER, 2 vs 7 vs 7 months, *P* = 0.003; for LR, 2 vs 13 vs 11 months, *P* = 0.005) (**S3 Figure** and **S4 Figure**).

**Figure 1.**
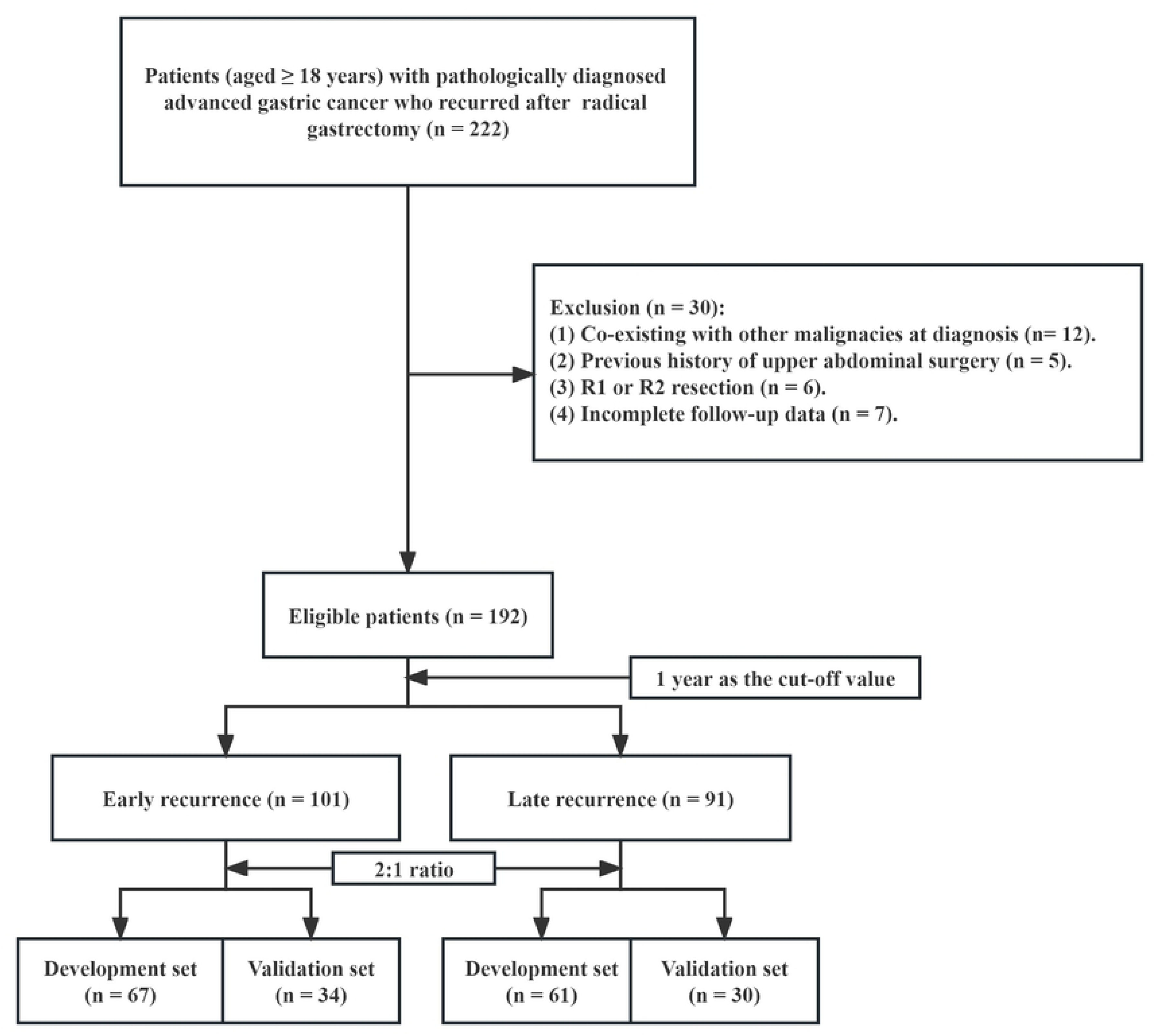
The flowchart of study population.

**Figure 2.**
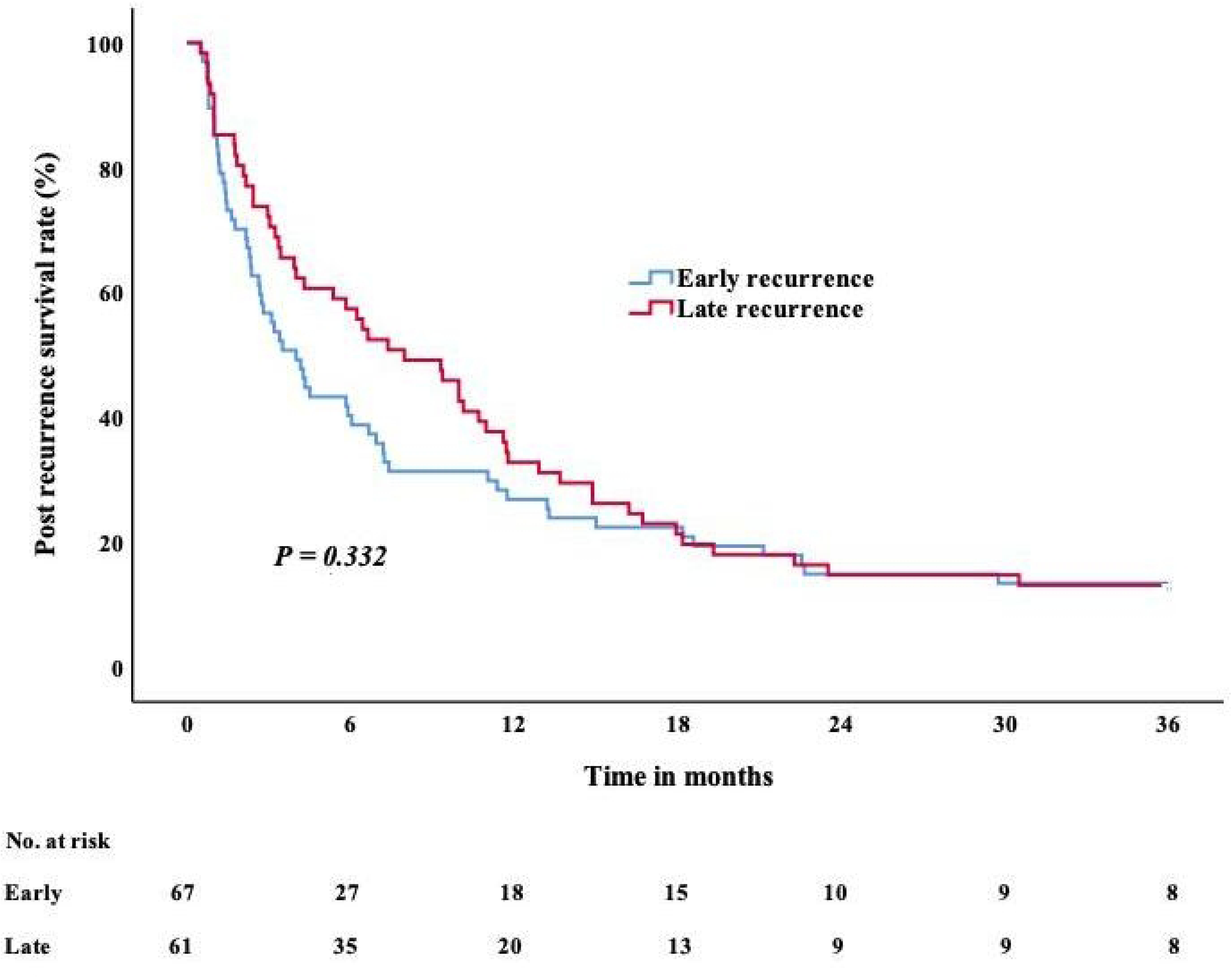
Kaplan-Meier curve of PRS for ER and LR. PRS, post-recurrence survival; ER, early recurrence; LR, late recurrence.

### Development and validation of nomograms

For patients with ER and LR, nomograms were constructed on the basis of these four independent predictors in the training set, respectively (**Figure 3, 4**).

**Figure 3.**
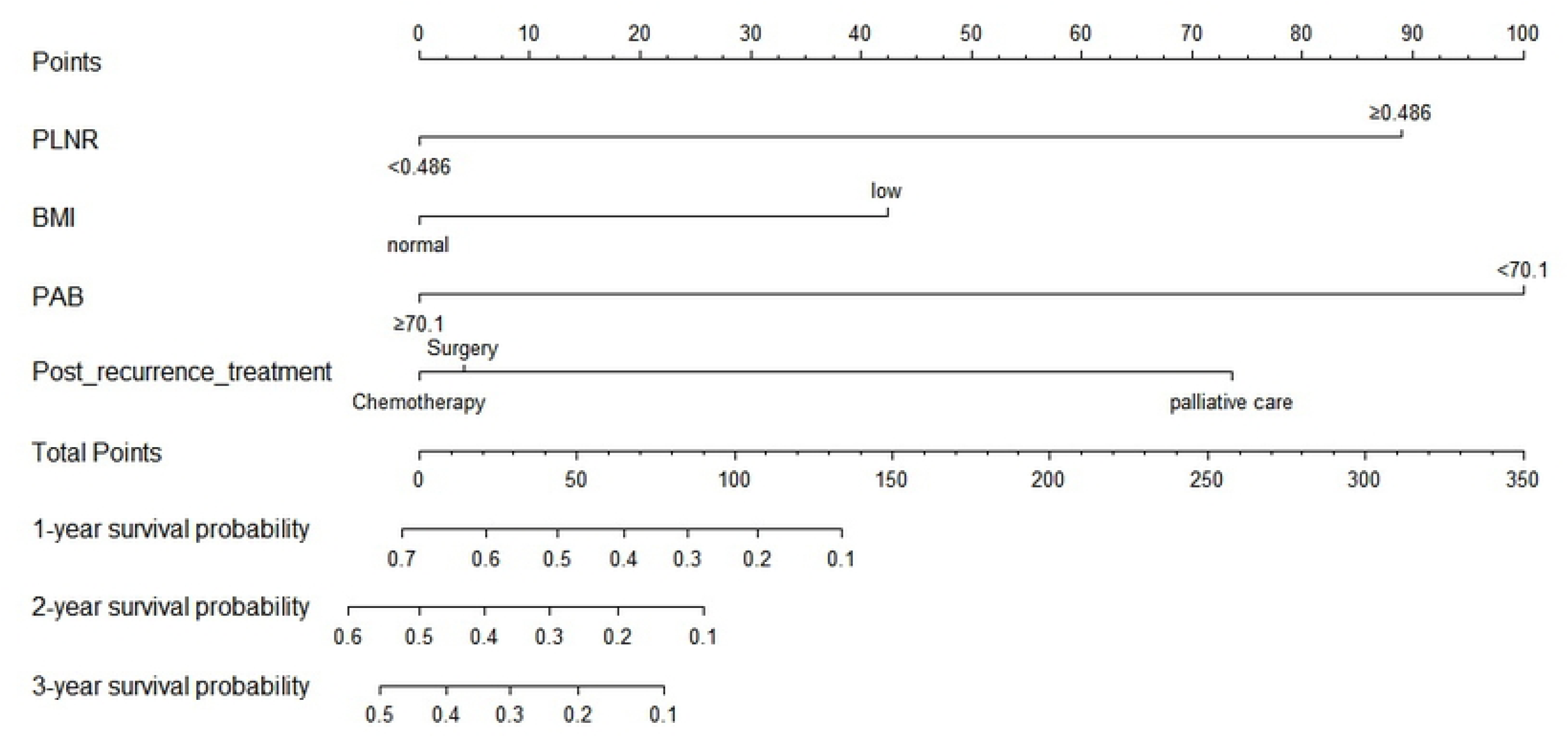
Nomogram including risk factors of PRS for ER. PRS, post-recurrence survival; ER, early recurrence.

**Figure 4.**
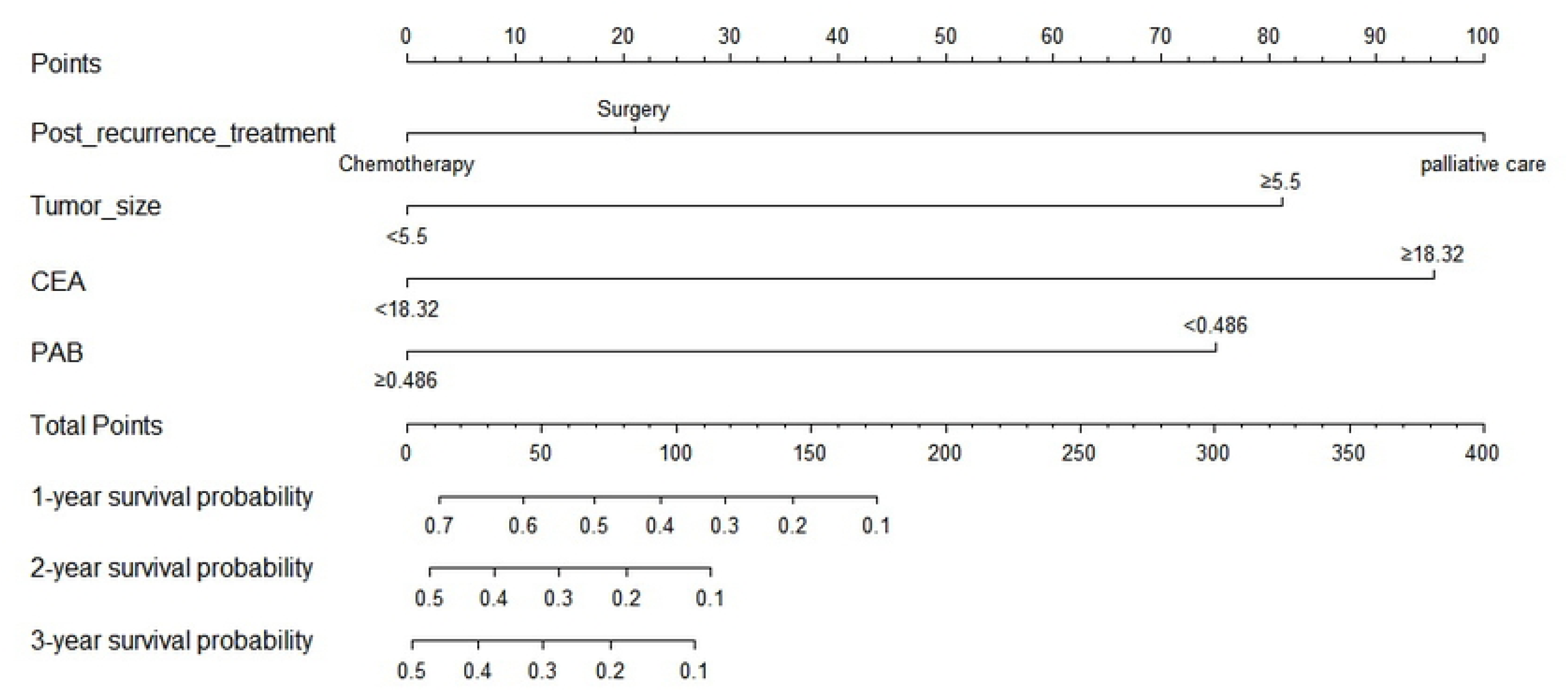
Nomogram including risk factors of PRS for LR. PRS, post-recurrence survival; LR, early recurrence.

For ER, the ROC curves of training and validation set showed that the AUC of the nomogram was 0.801 (95% CI, 0.746-0.856) and 0.744 (95% CI, 0.630-0.856), respectively (**Figure 5a, 5b**). The calibration curves showed a good fit between prediction and actual observation (**Figure 5c, 5d**). Then, the decision curves also demonstrated good predictive power (**Figure 5e, 5f**).

**Figure 5.**
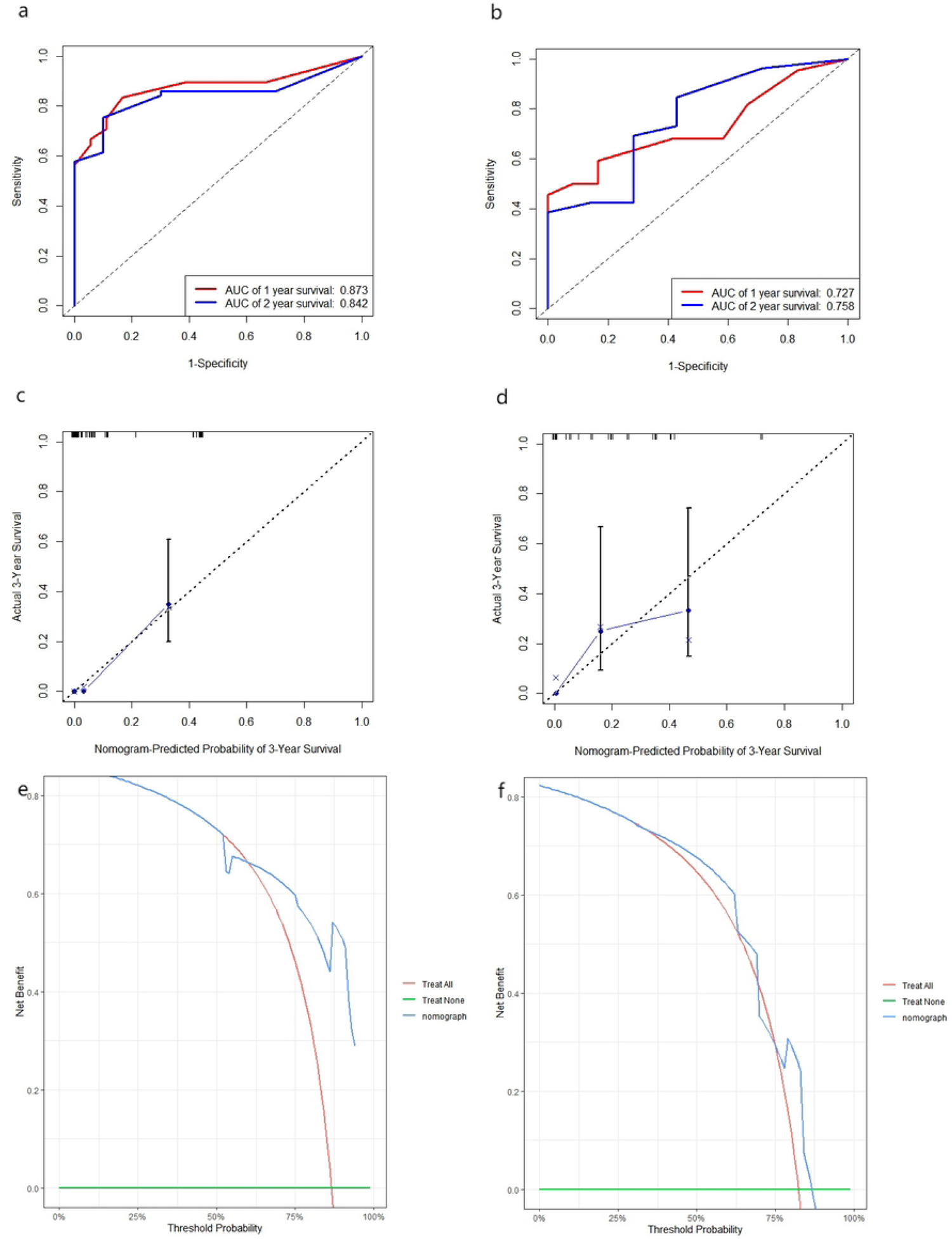
Receiver operating characteristics (**a**. training set; **b**. validation set), calibration curves (**c**. training set; **d**. validation set) and decision curves (**e**. training set; **f**. validation set) for predicting PRS of ER. PRS, post-recurrence survival; ER, early recurrence.

Similarly, for LR, the ROC curves of training and validation set showed that the AUC of the nomogram was 0.772 (95% CI, 0.709-0.835) and 0.676 (95% CI, 0.582-0.770), respectively (**Figure 6a, 6b**). The calibration curves showed a good fit between prediction and actual observation (**Figure 6c, 6d**). Then, the decision curves also demonstrated good predictive power (**Figure 6e, 6f**).

**Figure 6.**
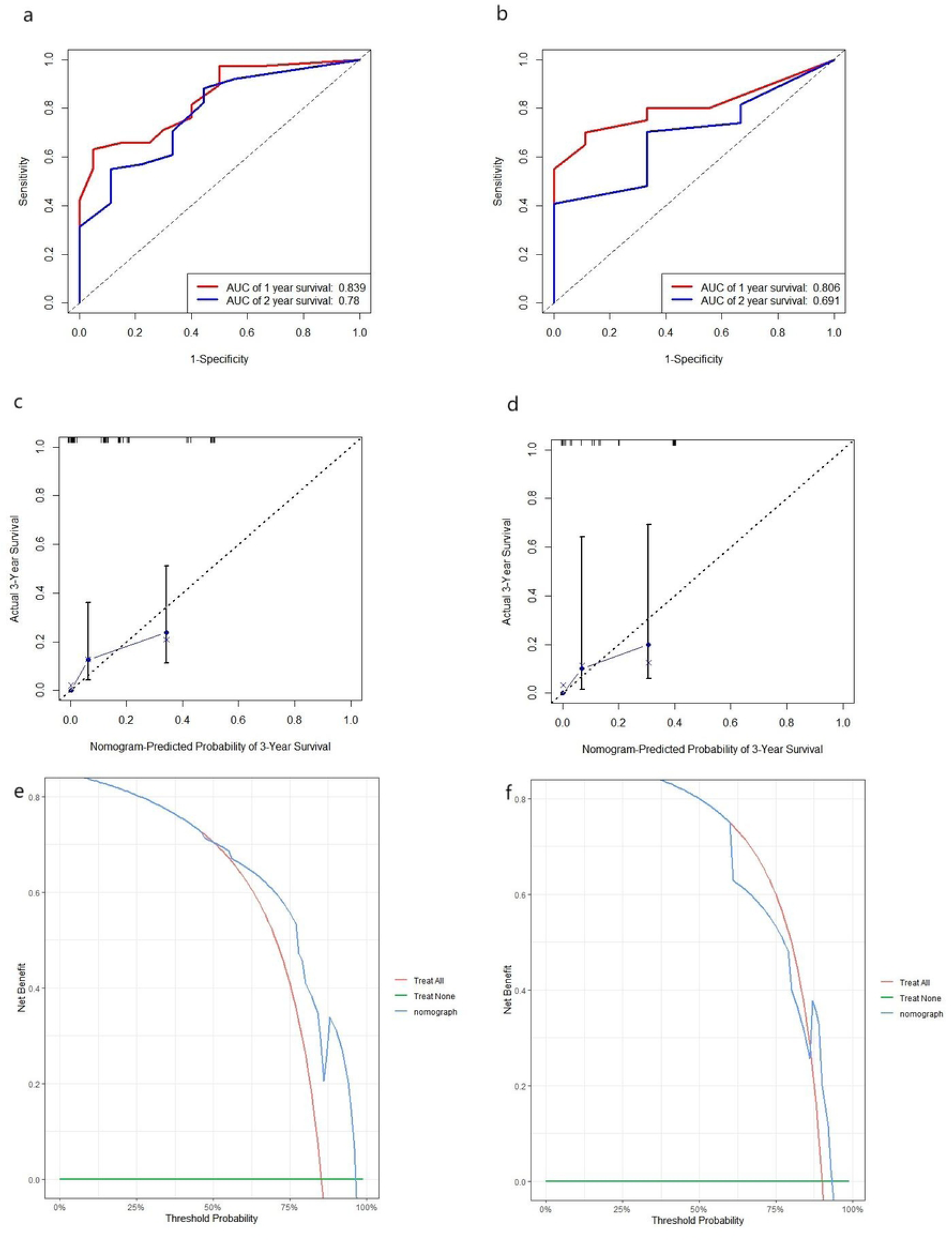
Receiver operating characteristics (**a**. training set; **b**. validation set), calibration curves (**c**. training set; **d**. validation set) and decision curves (**e**. training set; **f**. validation set) for predicting PRS of LR. PRS, post-recurrence survival; LR, late recurrence.

### Exploration of the predictive ability of nomogram

According to different nomogram scores, the predictive probability of ER was divided into two risk groups (low risk group and high risk group) to further evaluate the predictive power of the nomogram.

As shown in **Figure 7a, b**, Kaplan-Meier survival curves showed that the nomogram risk grouping had better discrimination power for PRS in the training and validation set (*P* < 0.001, *P* = 0.031). The median PRS in the low-risk group was significantly longer than that in the high-risk group for ER (11 months vs 1 month; 13 months vs 1 month).

**Figure 7.**
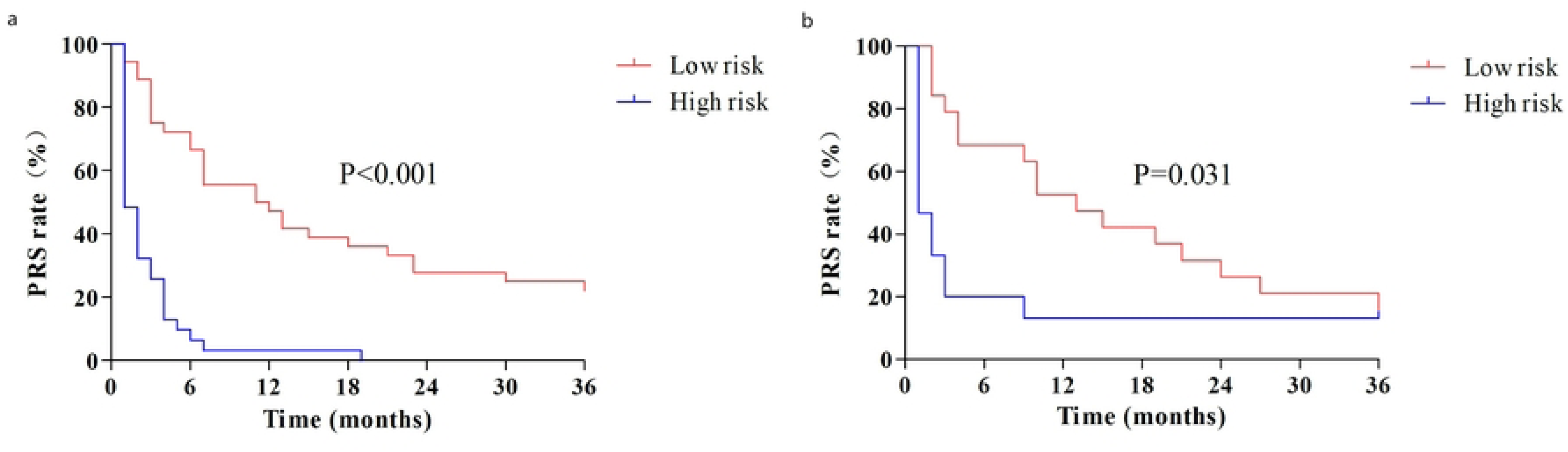
Kaplan-Meier curve of PRS between high-risk and low-risk group for ER. PRS, post-recurrence survival; ER, early recurrence. (**a**). training set; (**b**). validation set.

Similarly, for LR, Kaplan-Meier survival curves showed that the nomogram risk grouping had better discrimination power for PRS in the training and validation set (*P* < 0.001, *P* = 0.005). The median PRS in the low-risk group was significantly longer than that in the high-risk group (13 months vs 3 months; 14 months vs 3 months) (**Figure 8a, b**).

**Figure 8.**
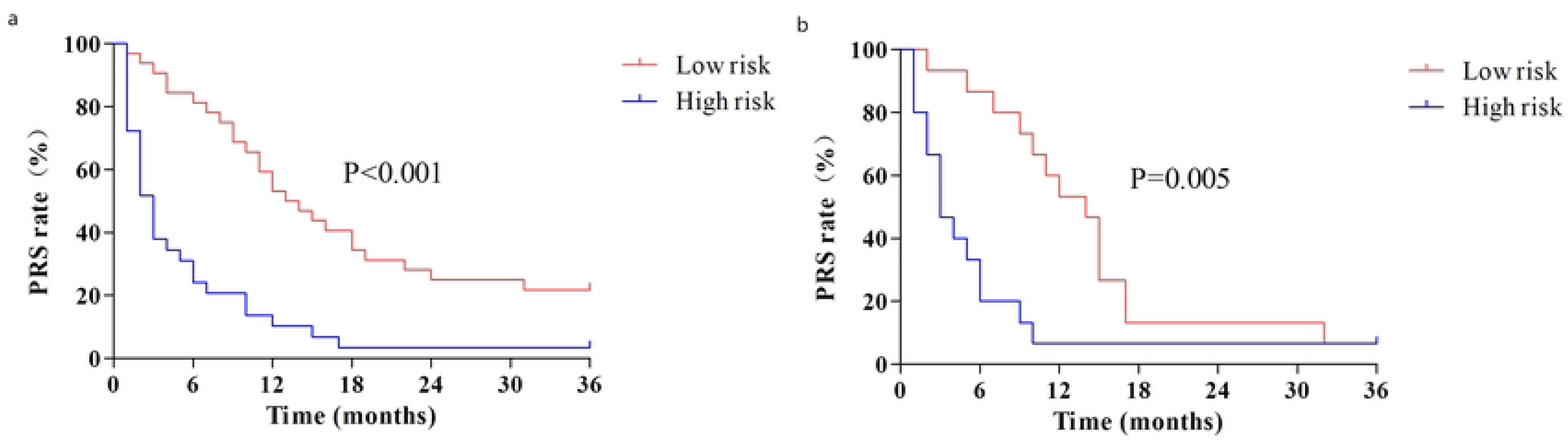
Kaplan-Meier curve of PRS between high-risk and low-risk group for LR. PRS, post-recurrence survival; LR, late recurrence. (**a**). training set; (**b**). validation set.

## Discussion

At present, the prognosis of AGC remains unsatisfactory, mainly due to its great possibility of recurrence and metastasis.^2^ Recurrence is a common and fatal condition in a variety of malignancies, including GC, and often indicates a poor prognosis.^6,7,8,9^ To the best of our knowledge, our study is the first to develop and validate nomograms in AGC patients with ER and LR, respectively. Our newly developed nomograms enable surgeons to accurately evaluate the prognosis of GC patients with ER and LR by stratification according to different risk factors, so as to take personalized treatment measures.

In this study, we revealed that there existed some differences in predictive factors of PRS between ER and LR. Low BMI and prealbumin level at the time of recurrence, high PLNR and palliative treatment after recurrence were risk factors for the prognosis of ER. In contrast, low prealbumin and high CEA level at recurrence, large tumor diameter and palliative treatment after recurrence were risk factors for the prognosis of LR.

Ma et al. first developed a nomogram for predicting ER of stage II/III GC after surgery based on tumor’s location, pTNM stage, lymphocyte count, postoperative complications, adjuvant chemotherapy and PLNR. Their C-index value was 0.780 with excellent practicability.^20^ Similarly, Liu et al. accurately predicted the role of age, Lauren classification, preoperative CA 19-9 level, pathological stage, major pathological response, and postoperative complications in early postoperative recurrence of GC.^11^ However, unlike these studies, we focused on the predictive role of indicators at and after recurrence. In the present study, for ER, the C-indexes were 0.801 and 0.744 in the training and validation cohort, respectively; and for LR, the C-indexes were 0.772 and 0.676, respectively. And the fit degree between the actual and predicted curve was in accord, indicating our prediction model had strong prediction power.

We found that high level of CEA at recurrence and large tumor size were independent risk factors for prognosis after LR. Previous studies have shown that patients with elevated CEA before surgery have an increased risk of recurrence, especially for ER.^21,22^ However, CEA levels at the time of recurrence in our study were only confirmed in LR. We hypothesized that this inconsistent phenomenon was related to the inclusion of indicators at two different periods, that was, before surgery and at recurrence. Although this has not been well proven, tumors may have different biological characteristics at recurrence than before surgery.

We observed a strong correlation between low BMI at recurrence and the prognosis of ER, but not in the cohort with LR. Consistent with our study, in a large study conducted in Chinese population to dynamically monitor the association between postoperative BMI and prognosis in GC patients, BMI loss of more than 10% within the first year after surgery was found to be associated with poor prognosis.^23^ We speculate that BMI is more reflective of changes in the nutritional status of the body in the early postoperative period.

In the present study, PLNR ≥ 0.486 was demonstrated an independent risk factor for survival of patients with ER, which was basically consistent with Ma et al., who found that patients with PLNR ≥ 0.335 had higher risk of ER than patients with PLNR ≤ 0.335 with stage II/III GC (OR: 3.37, 95% CI 2.37-4.78, *P* < 0.001).^20^ However, this relationship was not observed in patients with LR. Moreover, Komtasu et al. concluded that patients with PLNR ≥ 0.4 had higher rate of node recurrence than those with PLNR < 0.4 in pN3 GC.^24^ So we speculate early lymph node recurrence might explain the association between higher PLNR and ER. However, the association of PLNR with the prognosis of GC after recurrence has not been fully confirmed.

We found a high level of prealbumin demonstrated a positive association with the survival in GC patients who experienced recurrence by setting the cut-off value of 70.1mg/l for ER and 170.1 mg/l for LR based on ROC curve. This relevance was also observed in previous studies. Shen et al. evaluated the clinical significance of preoperative prealbumin in 731 stage II/III GC patients with a cut-off value of 180 mg/l.^25^ The median OS was 62 months in the high-prealbumin group and 46 months in the low-prealbumin group (HR: 1.362, 95% CI, 1.094–1.695). Aoyama et al. found that the patients with a prealbumin level < 20 mg/dl had significantly poorer outcomes than those with higher prealbumin levels.^26^ The HR for the OS was 2.375 (95% CI, 1.362–4.144). In contrast to prealbumin, a statistical association was not found between albumin and PRS. Consequently, owing to its shorter half-time than albumin, prealbumin is a more sensitive index of nutritional change than albumin.^27,28^ Our results suggest that if proper nutritional support before recurrence could be given, a low prealbumin concentration may serve as a modifiable risk factor for prognosis. Large scale randomized controlled trails should be conducted in the future.

Regarding the post-recurrence treatment strategies, we found that the prognosis of recurrent GC patients with re-surgical intervention and adjuvant chemotherapy was significantly better than that of those with palliative treatment alone. However, no significant difference was observed between re-surgical intervention and adjuvant chemotherapy only in median PRS. Compared with other malignant solid tumors, such as recurrent hepatocellular carcinoma and rectal adenocarcinoma, recurrent GC patients rarely received re-surgical intervention, only 14.9% and 21.3% of ER and LR patients, respectively in our study. Previous studies confirmed that re-surgical resection should be implemented only in selected patients with local recurrence in whom complete resection is possible.^29,30,31^

We have to admit that our study has several limitations. First, our study was a single-center retrospective study with limited data. The cut-off value for distinguishing ER and LR was selected based on previous studies, so there may be some deviation from the real data in this study. Second, the generalizability of our findings may be limited by the lack of external validation. Third, other important clinicopathological factors that have an important impact on the prognosis of recurrence, such as novel genetic markers, were not collected. Despite these limitations, our study is the first to develop predictive models based on independent risk factors for the prognosis of ER and LR of GC. In the future, expect external data from large-scale centers to validate the established models.

## Conclusion

In conclusion, Body mass index < 18.5 kg/m^2^, prealbumin level < 70.1 mg/l, Positive lymph nodes ratio ≥ 0.486 and palliative treatment after recurrence were independent risk factors for the prognosis of ER. In contrast, prealbumin level < 170.1 mg/l, CEA ≥ 18.32 μg/l, tumor diameter ≥ 5.5 cm and palliative treatment after recurrence were independent risk factors for the prognosis of LR. Nomograms based on these risk factors showed good predictive power and could provide reference for clinicians’ treatment strategies to some extent.

## Data Availability

All relevant data are within the manuscript and its Supporting Information files.

## Abbreviations

GC: gastric cancer
ER: early recurrence
LR: late recurrence
PNI: prognostic nutritional index
PLNR: positive lymph node ratio
ROC: receiver operating characteristic
BMI: body mass index
PRS: post-recurrence survival
SD: standard deviation
IQR: interquartile range
C-index: concordance index
CI: confidence interval
AUC: area under curve.

## Acknowledgement

We ’re thankful for the colleagues from the general surgery due to their generous assistance.

## Supporting information captions

S1 Table. The cut-off value for variables of early recurrence group by receiver operating characteristic.

S2 Table. The cut-off value for variables of late recurrence group by receiver operating characteristic.

S1 Figure. Kaplan-Meier curve of between different PAB levels for early recurrence group. PAB, prealbumin.

S2 Figure. Kaplan-Meier curve of between different PAB levels for late recurrence group. PAB, prealbumin.

S3 Figure. Kaplan-Meier curve of between different post-recurrence treatments for early recurrence group.

S4 Figure. Kaplan-Meier curve of between different post-recurrence treatments for late recurrence group.

